# The causal relationship between gastro-esophageal reflux disease and idiopathic pulmonary fibrosis: A bidirectional two-sample Mendelian randomization study

**DOI:** 10.1101/2022.08.31.22279411

**Authors:** Carl J Reynolds, Fabiola Del Greco M, Richard J Allen, Carlos Flores, R Gisli Jenkins, Toby M Maher, Philip L Molyneaux, Imre Noth, Justin M Oldham, Louise V Wain, Jiyuan An, Jue-Sheng Ong, Stuart MacGregor, Tom A. Yates, Paul Cullinan, Cosetta Minelli

## Abstract

**Background:** Gastro-esophageal reflux disease (GERD) is associated with idiopathic pulmonary fibrosis (IPF) in observational studies. It is not known if this association arises because GERD causes IPF, or IPF causes GERD, or because of confounding by factors, such as smoking, associated with both GERD and IPF. We used bidirectional Mendelian randomisation (MR), where genetic variants are used as instrumental variables to address issues of confounding and reverse causation, to examine how, if at all, GERD and IPF are causally related.

**Methods and results:** A bidirectional two-sample MR was performed to estimate the causal effect of GERD on IPF risk, and of IPF on GERD risk, using genetic data from the largest GERD (78,707 cases and 288,734 controls) and IPF (4,125 cases and 20,464 controls) genome-wide association meta-analyses currently available. GERD increased the risk of IPF, with an odds ratio (OR) of 1.6 (95% Confidence Interval, CI: 1.04-2.49; p=0.032). There was no evidence of a causal effect of IPF on the risk of GERD, with an OR of 0.99 (95%CI: 0.97-1.02; p=0.615).

**Conclusion:** We found that GERD increases the risk of IPF, but found no evidence that IPF increases the risk of GERD. GERD should be considered in future studies of IPF risk, and interest in it as a potential therapeutic target should be renewed. The mechanisms underlying the effect of GERD on IPF should also be investigated.

## Introduction

Idiopathic pulmonary fibrosis (IPF) is a progressive fibrotic lung disease characterised by usual interstitial pneumonia pattern on CT thorax or biopsy in the absence of a recognised cause. [1] The incidence in Europe and North America is 3-9 per 100,000 person-years [2] and the prognosis is poor, with a median survival of three years. [3,4] IPF is thought to result from epithelial injury in the distal airways, in a susceptible host, initiating a dysregulated repair process. [5] Environmental contributions to IPF are not well understood but several exposures have been posited as causal, these include smoking, diabetes mellitus, and gastro-esophageal reflux disease (GERD).

GERD is defined by non-physiological aspiration of gut contents associated with troublesome symptoms and/or complications such as esophagitis. [6] GERD has been associated with IPF in observational studies [7] but it is not known if GERD causes an increased risk of IPF. The observed association might be due to confounding by factors, such as smoking, associated with both GERD and IPF. The association could also result from reverse causation, with IPF causing an increased risk of GERD rather than vice-versa. [8] This is plausible given that reduced lung compliance in IPF can lead to more negative intrathoracic pressures and reflux events are inversely correlated with inspiratory thoracic pressures. [9, 10]

Unlike observational associations, genetic associations are not affected by classical confounding or reverse causation, as genes are randomly allocated at conception. A Mendelian randomisation (MR) approach that uses genetic variants known to affect GERD as its proxies (instrumental variables) can therefore provide indirect evidence for a causal effect of GERD on the risk of IPF, if its underlying assumptions hold. [11,12] Likewise, using genetic variants known to affect IPF risk as its proxies, the same approach can provide indirect evidence for a causal effect of IPF on the risk of GERD.

In this paper we describe a bidirectional, two-sample MR study to estimate the causal effect of GERD on the risk of IPF, and of IPF on GERD risk. We used data from the largest available genome-wide association meta-analyses for both GERD [13] and IPF. [14]

## Methods

### Genetic data

For both our analyses of the effect of GERD on IPF risk, and of the effect of IPF on GERD risk, we used two-sample MR where summary statistics (effect estimates and standard errors) for the gene-exposure (G-X) and gene-outcome (G-Y) associations were obtained from separate studies. A graphical overview of the two MR analyses is provided in Figure 1.

**Figure 1:**
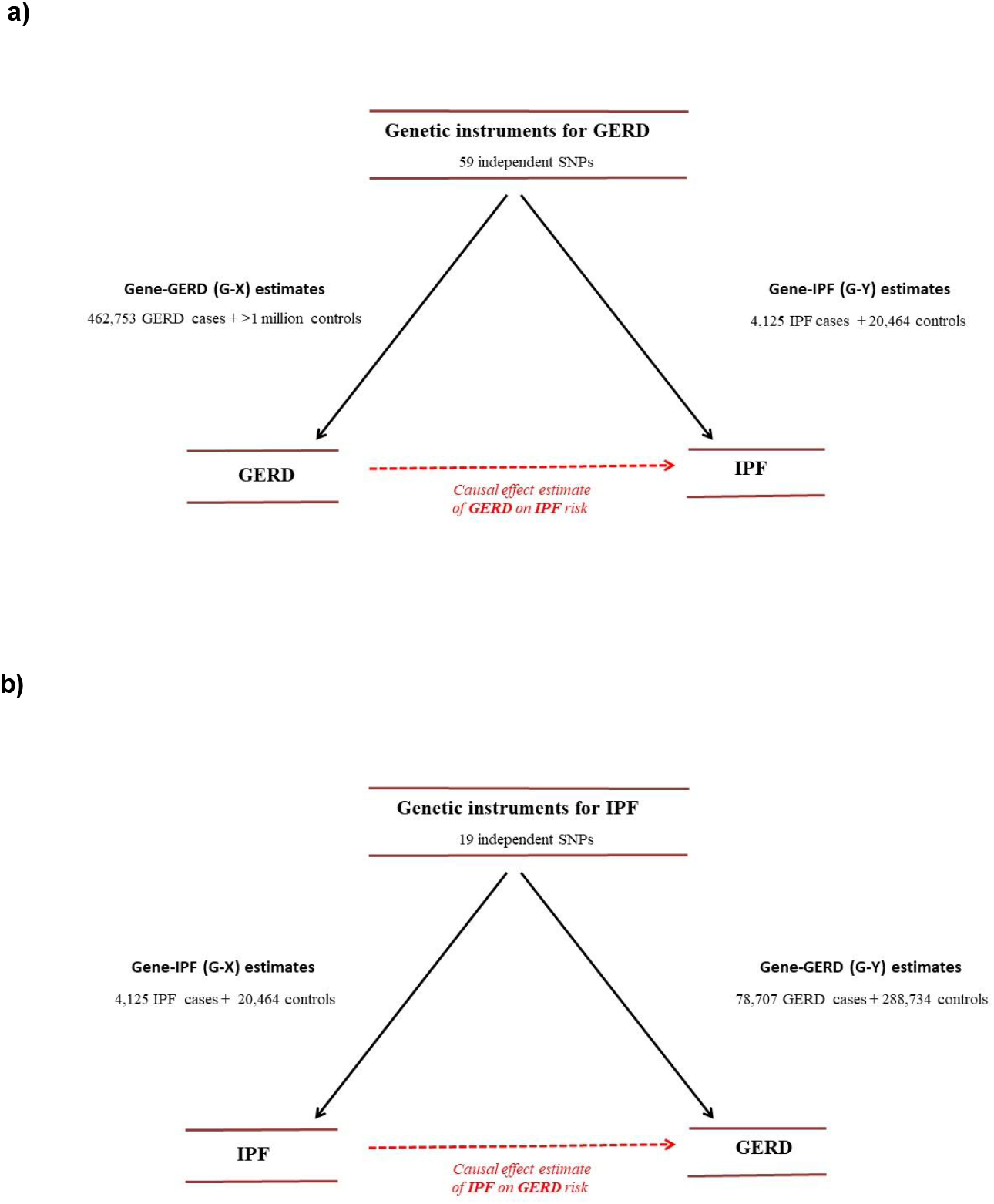
Overview of our two-sample MR analysis of: **a)** GERD on IPF risk; **b)** IPF on GERD risk. G-X: gene-exposure association; G-Y: gene-outcome association.

For the MR of the effect of GERD on IPF risk, instruments were selected from the largest available genome-wide association study (GWAS) meta-analysis on GERD by Ong et al. [13] For each instrument (Single Nucleotide Polymorphism, SNP), summary statistics of the G-X association (expressed as log OR for GERD) were obtained from the replication stage of Ong et al. [13] Summary statistics of the G-Y association (log OR for IPF) were obtained from the authors of the GWAS meta-analysis on IPF. [14]

Similarly, for the MR of the effect of IPF on GERD risk, instruments were selected from the largest available GWAS meta-analysis on IPF by Allen et al. [14]. For each SNP, summary statistics of the G-X association (log OR for IPF) were obtained from this GWAS, while summary statistics of the G-Y association (log OR for GERD) were obtained from the authors of the GWAS meta-analysis on GERD. [13]

### MR methods

Here we provide an overview of the MR methods used, with a more detailed description of these methods, their underlying assumptions, relevant references and the code used for the analyses reported in Supplementary Methods.

We estimated the causal effect of GERD on IPF risk (Figure 1a) and of IPF on GERD risk (Figure 1b) by first deriving SNP-specific MR estimates using the Wald estimator (G-Y divided by G-X), and then pooling them using inverse-variance weighted, fixed-effect meta-analysis (IVW-FE).

We used the IVW-FE method for our main MR analyses as this is the most powerful one, but it assumes absence of pleiotropy, i.e. variants chosen as instruments for the exposure cannot affect the outcome through any other independent pathways. Pleiotropy can bias MR findings, and we therefore investigated its possible presence through assessment of the heterogeneity in the MR estimates across SNPs, using the I^2^ index and Cochran’s Q heterogeneity test.

In the presence of pleiotropy, possible pleiotropic SNPs were identified graphically based on their contribution to the overall heterogeneity (Cochran’s Q statistic), and we repeated the IVW-FE analysis after removing the pleiotropic SNPs. We also performed MR analyses on all SNPs using methods that can account for pleiotropy under different assumptions about its nature. In particular, we considered the following methods: inverse variance weighted random-effect (IVW-RE); Weighted Median (WMe); Weighted Mode-based (WMo); and MR-Egger (MRE), with the simulation extrapolation (SIMEX) method to correct for measurement error (dilution bias) used if needed.

## Results

Demographic data for the cohorts used is provided in Table 1 and described below.

**Table 1:** Summary information for the studies contributing to the data used in both MR analyses.

| Study |  | N | Age<br>(Mean (SD)) | Sex<br>(% Males) | Ever smokers<br>% | Genotyping array | Imputation<br>panel |
| --- | --- | --- | --- | --- | --- | --- | --- |
| GERD |  |  |  |  |  |  |  |
| UK | Cases | 75720 | 59 | — | — | Affymetrix UK BiLEVE array | HRC |
|  | Controls | 278565 | 57 | 46 | 41 |  |  |
| Australia | Cases | 2987 | 56.5 | — | — | Illumina Global Screening<br>Array | HRC |
|  | Controls | 10169 | 56 | 46 | 54 |  |  |
| IPF |  |  |  |  |  |  |  |
| Chicago | Cases | 541 | 68(3) | 71 | 72 | Affymetrix 6.0 SNP array | HRC |
|  | Controls | 542 | 63(7.5) | 47 | 42 |  |  |
| Colorado | Cases | 1515 | 66(9.5) | 68 | — | Illumina Human 660W Quad<br>BeadChip | HRC |
|  | Controls | 4683 | — | 49 | — |  |  |
| UK | Cases | 612 | 70(8.4) | 71 | 73 | Affymetrix UK BiLEVE array | HRC |
|  | Controls | 3366 | 65(5.5) | 70 | 70 | Affymetrix UK BiLEVE and UK<br>Biobank arrays |  |
| UUS | Cases | 793 | 69(8.1) | 75 | 69 | Affymetrix UK Biobank and<br>Spain Biobank arrays | HRC |
|  | Controls | 9999 | 58(7.8) | 72 | 68 | Affymetrix UK BiLEVE and UK<br>Biobank arrays |  |
| Genentech | Cases | 664 | 68(7.5) | 74 | 67 | HiSeq X Ten platform<br>(Illumina) |  |
|  | Controls | 1874 | 56(9.3) | 27 | 18 |  |  |

### Effect of GERD on IPF risk

The GWAS on GERD by Ong et al. [13] identified 59 independent genome-wide significant (p<5×10^−8^) SNPs, based on a total sample of 78,707 GERD cases and 288,734 controls, with replication of findings in 462,753 cases and 1,484,025 controls. [13] The sample was almost entirely of White European ancestry. GERD cases were defined by having one or more of a GERD ICD10 code, GERD self-report, or use of GERD medication.

Of the 59 SNPs, two were missing in the IPF GWAS meta-analysis dataset and a proxy (i.e. SNP with linkage disequilibrium, LD, r^2^≥0.8 with the SNP of interest) was used instead (Supplementary Table 1). All SNPs used in the MR analysis were “strong” instruments with an F statistic>10 [15], where the F statistic is a function of both magnitude and precision of the SNP’s effect on GERD. [16] Individual F statistics ranged from 12 to 81 (Supplementary Table 1).

This MR analysis showed that GERD increases the risk of IPF, with an odds ratio (OR) of 1.61 (95% Confidence Interval, CI: 1.04-2.49; p=0.032).

There was no statistical evidence of pleiotropy, with an I^2^ of 0% (95%CI: 0-31%), and, a heterogeneity p-value of 0.52.

### Effect of IPF on GERD risk

The GWAS meta-analysis on IPF by Allen et al. [14] included three previously published studies (Chicago, Colorado, UK) [17–19] plus an unpublished study including independent case-control studies from the US, UK, and Spain (UUS), and a study of clinical trial subjects, the Genentech study. The Genentech study consisted of cases from three IPF clinical trials and controls from four non-IPF clinical trials.

The GWAS meta-analysis included a total of 4,125 IPF cases and 20,464 controls of White European ancestry. IPF cases were defined using the joint American Thoracic Society and European Respiratory Society guidelines. [20] [21] [1]

Of the 19 SNPs, 15 were missing in the GERD GWAS meta-analysis database. For nine a proxy (LD r^2^>0.8) was used (Supplementary Table 2). Data for the remaining six, for which a satisfactory proxy could not be identified, were obtained from the authors. Individual F statistics ranged from 30 to 1,721 (Supplementary Table 2).

The MR analysis showed no evidence of a causal effect of IPF on the risk of GERD, with an OR of 0.999 (95%CI: 0.997-1.001; p=0.245).

We found evidence of pleiotropy, with an I^2^ of 63% (95%CI: 40%-77%), and a heterogeneity p-value of 0.00016. We identified 5 possible pleiotropic SNPs, shown in Figure 3. After removing them, there was no evidence of residual pleiotropy (I2 of 0%; heterogeneity p-value of 0.944), and the results remained null (OR of 0.998; 95%CI: 0.996-1.001; p=0.137). Similar null findings were obtained when using robust methods that adjust for pleiotropy, as shown in Figure 2.

**Figure 2:**
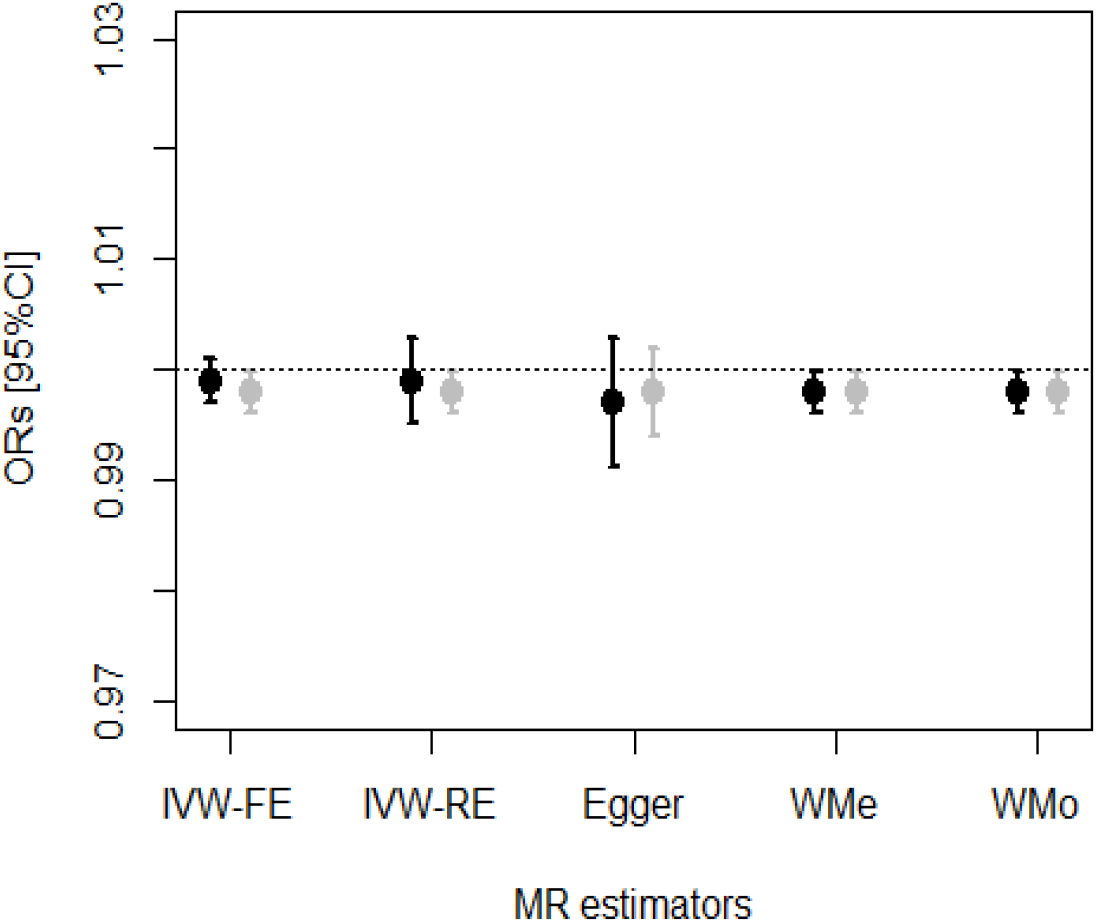
Results of the MR on the effect of IPF on GERD risk using different robust methods to address the issue of pleiotropy. Black for main analysis, grey for sensitivity analysis with five possible pleiotropic SNPs removed.

**Figure 3:**
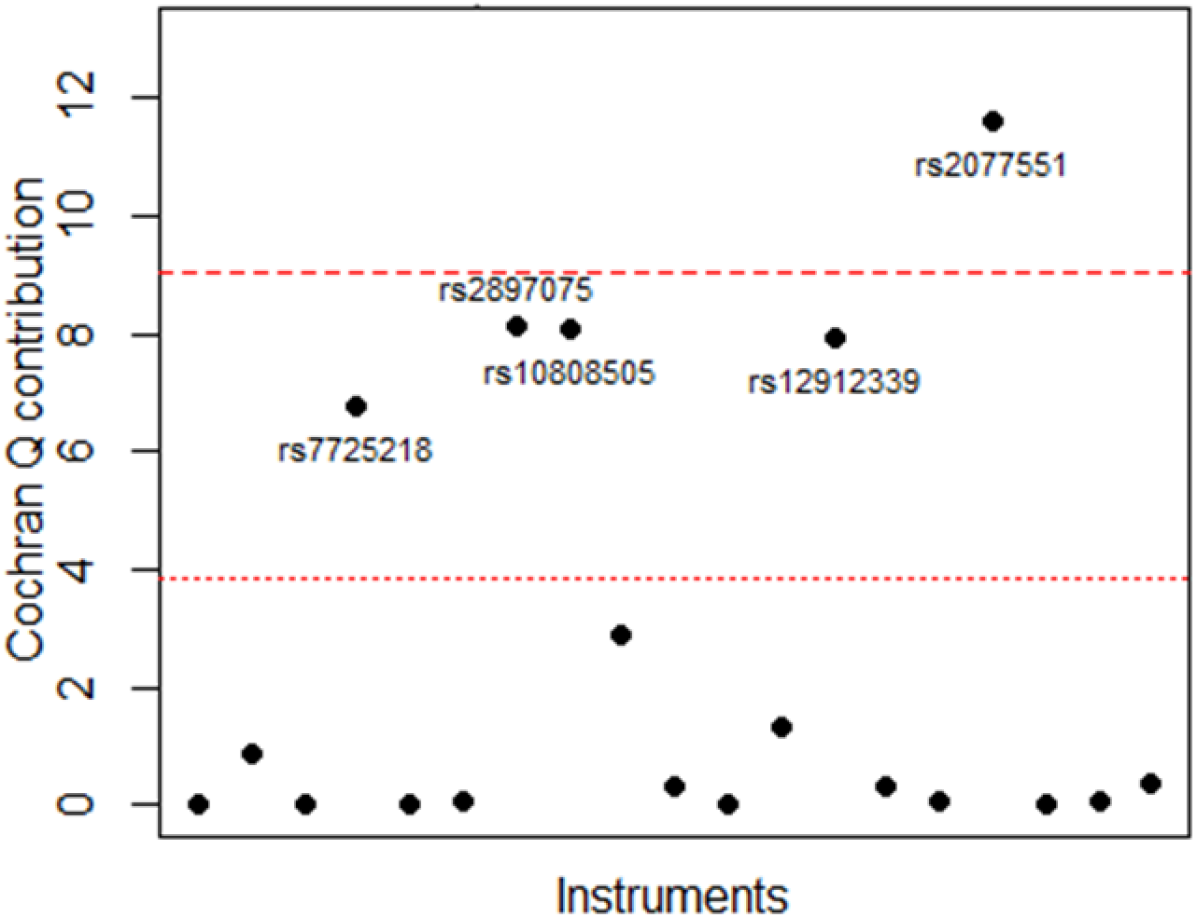
Identification of pleiotropic instruments in the MR on the effect of IPF on GERD. Individual variant contributions to Cochran’s Q heterogeneity statistic with the 5^th^ (dotted line) and Bonferroni corrected (0.05/19^th^) percentiles (dashed line) of a chi-squared with 1 degree of freedom.

## Discussion

We undertook a bidirectional two-sample MR study to investigate the causal relationship between GERD and IPF. We found evidence of a causal effect of GERD on IPF, but not of IPF on GERD.

The odds of IPF were 1.6 times higher in the presence of GERD (95%CI: 1.04-2.49; p=0.032). Previous evidence for the association of GERD and IPF arises from observational studies where it has not been possible to confidently make causal inferences because of the potential for residual confounding and reverse causation. A case-control study of 17 consecutive biopsy-proven IPF patients and eight controls with non-IPF ILD at a US tertiary ILD centre [22], which involved ambulatory esophageal pH monitoring and a GERD symptom questionnaire, found that the majority of IPF, but not non-IPF ILD, patients had abnormal esophageal acid exposure which was usually clinically silent. A similar pattern was seen in a later study of 65 consecutive IPF patients at the same centre, which used 133 consecutive asthma patients referred to a gastrointestinal motility clinic as controls. [23] A systematic review of IPF comorbidities found 23 studies that reported prevalence of GERD in IPF patients. The majority of the studies were small, with fewer than 50 patients, and they used a variety of case definitions for GERD. [24] Study estimates varied widely, from 0 to 94%, but most were around 30%. [24] This is higher than prevalence estimates for the general population; GERD global population prevalence increases with age and peaks at 18% for people aged 75-79. [25] A recent meta-analysis of 18 case-control studies [7] comprising 3206 patients with IPF and 9368 controls, found the odds of IPF was 2.94 time higher in people with GERD (95% CI: 1.95-4.42), but with high heterogeneity across studies (I^2^ = 86%; heterogeneity p-value<0.00001). The authors reported that confounding by smoking was likely for two reasons. First, a post-hoc meta-regression that controlled for smoking found the association was no longer significant (OR: 0.66; 95% CI: 0.34-1.27). Second, the effect sizes tended to be larger in studies where a higher proportion of cases, and a lower proportion of controls, smoked. There was significant correlation between the ratio of the proportion of smokers (and ex-smokers) in IPF cases over the proportion in controls and the log odds ratio for the association of GERD and IPF. Our findings using an MR approach overcome confounding by smoking and any other (known or unknown) confounding of observational analyses to indicate a causal effect of GERD on IPF risk.

The underlying mechanism explaining how GERD may increase IPF risk is unknown, however, aspiration of gastric contents can cause chemical pneumonitis or aspiration pneumonia. [26] One study of GERD in IPF included bronchoalveolar lavage (BAL) and compared 21 IPF patients with 20 non-IPF ILD patients and 16 patients undergoing bronchoscopy for other diseases. BAL fluid was analysed for the presence of pepsin and bile acids. Pepsin and/or bile acids were present in 62% of IPF patients compared with 25% of non-IPF ILD patients, and were absent for all patients undergoing bronchoscopy for other diseases. [27]

While direct chemical or bacterial epithelial insult may initiate or promote fibrosis another possibility is by indirect means. For example, GERD may increase IPF risk through airway acidification secondary to aspiration disrupting MUC5B function and impairing innate immunity. The *MUC5B* promoter variant rs35705950, which increases airway expression of MUC5B, is the strongest common identified risk factor for IPF. The odds of developing pulmonary fibrosis are 5 times higher for individuals carrying one copy of the disease allele, rising to 20 times higher for individuals with two copies, when compared with individuals carrying no copies of the disease allele. The frequency of the disease allele is around 9% in European ancestry populations. [28,29] While MUC5B is undoubtedly important for IPF risk, the mechanisms by which increased airway MUC5b expression increases IPF risk are not well understood. In evolutionary terms, it is a highly conserved airway mucin that plays a key role in antimicrobial host defence [30] and has been shown to reduce mucosal bacterial load and to promote mucosal microbial diversity. [31] The structure, and function, of MUC5B is pH dependent [32] [33] and may be disrupted by acidification of the airway secondary to GERD. In IPF, there are higher airway bacterial loads and reduced microbial diversity compared with COPD patients and healthy controls [34] and this is associated with disease progression. [35]

The theoretical possibility of reverse causation, whereby the association between IPF and GERD is driven by IPF, rather than GERD, is well described. [36] Restrictive lung disease may distort the esophageal gastric junction and predispose to hiatus hernia. Indeed, hiatus hernia has been observed to be more common in IPF than in patients with asthma or COPD. [37] Reduced lung compliance in IPF can result in increasingly negative intrathoracic pressures being required for inspiration and an increased gastro-oesophageal pressure gradient. [38] Reflux events generally occur during inspiration and are inversely correlated with inspiratory thoracic pressures. [9] [10] Other mechanical factors that might contribute to GERD secondary to IPF are reduced esophageal body motility, lower basal lower esophageal sphincter pressure, and delayed gastric emptying. [36] We therefore investigated the possibility of causality in both directions using a bidirectional MR approach, but we found no evidence of a causal effect of IPF on GERD risk. In this MR analysis of the effect of IPF on GERD, however, pleiotropy was observed and five possible pleiotropic SNPs identified, but the result remained null after removing them and when adjusting for pleiotropy using robust methods.

A major strength of our MR work, in contrast to previous observational work examining GERD-IPF relations, is that MR is not affected by classical confounders, such as smoking, or by reverse causation. Our work therefore overcomes these limitations of observational studies to establish GERD as a risk factor for IPF.

Our study also has some limitations. One major problem of MR is that while it is not vulnerable to classical confounding or reverse causation it is affected by pleiotropy. We found evidence for pleiotropy in our MR of IPF on GERD but adjusting for it did not alter our results. Our study had more power for finding an effect of GERD on IPF than IPF on GERD as a consequence of GERD being more common, and there being many more genotyped GERD patients than IPF patients available. We cannot exclude that a small effect of IPF on GERD might be found in a future MR study performed on data from a larger IPF sample. Finally, participants contributing to the GWAS meta-analyses used were almost entirely of White European ancestry, limiting generalisability to other groups.

GERD should be taken into account in future studies of IPF risk and interest in it as a potential therapeutic target should be renewed. Treatment of GERD is not well established to be of benefit in IPF; a recent systematic review found no evidence that treating GERD with antacids, or fundoplication, improved outcomes in IPF. [39] There is also potential for harm, since adverse events including increased risk of bacterial infection have been associated with antacid use. [39] Consequently, international guidelines have been updated to include a conditional recommendation against, rather than for, antacid treatment in IPF. Uncertainty remains due to a lack of adequately powered well designed randomised controlled trials (RCT) in this area, though one is now underway. [40]

We have assessed the risk of developing IPF in GERD, which is not amenable to indirect assessment by an RCT of GERD treatment, given how rare IPF is an outcome. It could be that GERD has also an independent effect on IPF disease progression; this could be investigated in a future study when sufficient genetic data becomes available. Understanding the mechanism for increased IPF risk in GERD is now necessary to understand which patients may benefit from existing treatments, and for identification of novel therapeutic targets.

## Conclusion

By means of a bidirectional two-sample MR study we have overcome limitations inherent to observational studies and shown that GERD causes an increased risk of IPF, while we found no evidence that IPF causes an increased risk of GERD. GERD should be considered in future studies of IPF risk, and interest in it as a potential therapeutic target should be renewed. The mechanisms underlying the effect of GERD on IPF should also be investigated.

## Data Availability

All data produced in the present study are available upon reasonable request to the authors

