## Supplementary Methods for "The causal relationship between gastro-esophageal reflux disease and idiopathic pulmonary fibrosis: A bidirectional two-sample Mendelian randomization study"

*MR methods*

We estimated the causal effect of GERD on IPF risk and of IPF on GERD risk by first deriving SNP-specific MR estimates using the Wald estimator, obtained by dividing the gene-outcome (G-Y) by the gene-exposure (G-X) association estimate (both expressed as logOR), with standard error obtained using the Delta method. [1] The estimate of the causal effect was then obtained by pooling the SNP-specific MR estimates using an Inverse-Variance Weighted Fixed-Effect meta-analysis (IVW-FE). [1] Although the linearity assumption underlying our MR analysis is violated when using binary variables [2], as in our case, the magnitude of the resulting bias has been shown to be negligible. [3]

*Investigation of pleiotropy*

The IVW-FE method used for our MR analyses is the most powerful but assumes absence of pleiotropy [4], i.e. variants chosen as instruments for the exposure cannot affect the outcome through any other independent pathways. As pleiotropy can bias MR findings [5], we investigated its possible presence through assessment of: the heterogeneity in the MR estimates across SNPs, using the $I^2$ index and the Q heterogeneity test [6].

In the presence of pleiotropy, possible pleiotropic SNPs were identified graphically based on their contribution to the overall heterogeneity (Cochran's Q statistic) as previously described [7], and we repeated the IVW-FE analysis after removing the pleiotropic SNPs. We also repeated the MR analyses on all SNPs using methods that can account for pleiotropy under different assumptions about its nature. [8] In particular, we considered the following methods:

- Inverse Variance Weighted Random-Effect (IVW-RE)[9]: This was performed in the same way as IVW-FE, but a (multiplicative) *random-effects*, instead of a *fixed-effect*, meta-analysis model was used to allow for pleiotropy. IVW-RE assumes that pleiotropic effects across SNPs are random (balanced pleiotropy), and that their magnitude is independent of the magnitude of the corresponding G-X effects (InSIDE assumption). [4]
- Weighted Median estimator (WMe) [10]: This method assumes that more than 50% of the information contributing to the MR analysis comes from genetic variants that are valid (i.e. they are not pleiotropic). [10]
- Weighted Mode estimator (WMo) [11]: This method assumes that the largest weighted contribution of "similar" (i.e. identical in infinite samples) SNP-specific MR estimates comes from valid instruments. [12]
- MR-Egger regression (MRE) [13]: In this method, G-Y estimates for the individual SNPs are regressed on their G-X estimates; the intercept of this regression model represents the overall pleiotropy, and the slope the MR estimate adjusted for pleiotropy .[14] MRE assumes an overall directional pleiotropy, and it makes the InSIDE assumption. MRE works well only in the presence of a large spread of strengths, which can be quantified by the heterogeneity in G-X estimates across SNPs, $I^2_{GX}$, with a recommended $I^2_{GX}$ > 90%. [13] As the $I^2_{GX}$ was lower than 90% for

both our MR analyses, we attempted to address this limitation using the SIMulation EXtrapolation (SIMEX) method that corrects for the dilution bias. [13]

The MR analyses were performed using the R packages “MendelianRandomization” (https://cran.r-project.org/web/packages/MendelianRandomization/index.html) and MR-PRESSO (https://github.com/rondolab/MR-PRESSO). The code for all the MR analyses is provided below.

*R code*

```
# Bi-directional Mendelian randomization analyses
# GERD and IPF

rm(list=ls())
#install.packages("MendelianRandomization")
#install.packages("simex")
#install.packages("MRPRESSO")
library(MendelianRandomization)
library(simex)
library(MRPRESSO)

A <- read.table("data.txt", sep="\t", header=T)
head(A)
dim(A)

#############################################
# MR-PRESSO
#############################################
mr_presso(BetaOutcome = "b_y", BetaExposure = "b_x", SdOutcome = "se_y", SdExposure
= "se_x", OUTLIERtest = TRUE, DISTORTIONtest = TRUE, data = A, NbDistribution = 1000,
SignifThreshold = 0.05)

#############################################
# IVW fixed effects
#############################################
mr_ivw(mr_input(bx = A$b_x, bxse = A$se_x, by = A$b_y, byse = A$se_y), model="fixed")

#############################################
#IVW multiplicative random effects
#############################################
mr_ivw(mr_input(bx = A$b_x, bxse = A$se_x, by = A$b_y, byse = A$se_y),
model="random")

##############################################
# MR-Egger
#############################################
```

```
mr_egger(mr_input(bx = A$b_x, bxse = A$se_x, by = A$b_y, byse = A$se_y))

##############################################
# MR-Egger with simex adjustment
##############################################
# dilution bias evaluation
##############################################
Isq <- function(y,s){
  k <- length(y)
  w <- 1/s^2; sum.w <- sum(w)
  mu.hat <- sum(y*w)/sum.w
  Q <- sum(w*(y-mu.hat)^2)
  Isq <- (Q - (k-1))/Q
  Isq <- max(0, Isq)
  return(Isq)
}

I2_gx <- Isq(A$b_x, A$se_x)
I2_gx

bxg <- A$b_x
seX <- A$se_x
byg <- A$b_y
seY <- A$se_y

BetaYG <- byg*sign(bxg)
BetaXG <- abs(bxg)

Fit2 <- lm(BetaYG~BetaXG,weights=1/seY^2,x=TRUE,y=TRUE)
mod.sim <- simex(Fit2,B=1000, measurement.error = seX,
SIMEXvariable="BetaXG",fitting.method ="quad",asymptotic="FALSE")
summary(mod.sim)

##############################################
# Weighted MR median
#############################################
mr_median(mr_input(bx = A$b_x, bxse = A$se_x, by = A$b_y, byse = A$se_y),
        weighting = "weighted", iterations = 10000)

##############################################
# Mode-based MR
##############################################
mr_mbe(mr_input(bx = A$b_x, bxse = A$se_x, by = A$b_y, byse = A$se_y), phi=1,
iterations=100)
QA1

#################################################################
# Plot of individual SNP contributions to Cochran's Q heterogeneity statistics
```

```
##################################################################

BIV <- A$b_y/A$b_x
se_IV <-sqrt(A$se_y^2/A$b_x^2)

w <- 1/(se_IV^2)
y <- BIV

sum.w <- sum(w)
mu.hat <- sum(y*w)/sum.w
Q_ivw <- w*(y-mu.hat)^2
su <- summary(Q_ivw)

plot(Q_ivw, pch = 19, ylab="", xlab = "", ylim=c(0,su[6]+1), xaxt = 'n', main="IPF-GERD")
title(ylab="Cochran Q contribution", line = 1.9)
title(xlab = "Instruments", line = 0.5)
L1 <- qchisq(1-0.05, df =  1)
L2 <- qchisq(1-(0.05/19), df =  1)

#add dotted line at 0.05
abline(L1,0,lty = 3, col = "red")

#add dotted line at 0.05/19
abline(L2,0, lty = 2, col = "red")

#Add rs names of instruments
text(4, Q_ivw[4], labels=A[4,1], cex= 0.6, pos=1)
text(7, Q_ivw[7], labels=A[7,1], cex= 0.6, pos=3)
text(8, Q_ivw[8], labels=A[8,1], cex= 0.6, pos=1)
text(13, Q_ivw[13], labels=A[13,1], cex= 0.6, pos=1)
text(16, Q_ivw[16], labels=A[16,1], cex= 0.6, pos=1)

##################################################################
```

4. Bowden J, Del Greco M F, Minelli C, Smith GD, Sheehan N, Thompson J. A framework

for the investigation of pleiotropy in two-sample summary data Mendelian randomization. Statistics in Medicine. 2017. pp. 1783–1802. doi:10.1002/sim.7221

5. Sheehan NA, Didelez V, Burton PR, Tobin MD. Mendelian Randomisation and Causal Inference in Observational Epidemiology. PLoS Medicine. 2008. p. e177. doi:10.1371/journal.pmed.0050177

6. Greco M FD, Minelli C, Sheehan NA, Thompson JR. Detecting pleiotropy in Mendelian randomisation studies with summary data and a continuous outcome. Stat Med. 2015;34: 2926–2940.

7. Bowden J, Del Greco M F, Minelli C, Zhao Q, Lawlor DA, Sheehan NA, et al. Improving the accuracy of two-sample summary data Mendelian randomization: moving beyond the NOME assumption. doi:10.1101/159442
